# Closed-Loop rTMS Induces Frequency-Specific Cortical Network Reorganization Distinct from Open-Loop Stimulation in Healthy Humans

**DOI:** 10.64898/2026.07.11.26357834

**Authors:** Federico Carparelli, Clara Simonetta, Davide Mascioli, Valerio Ferrari, Silvio Bagetta, Carlo Conti, Diego Centonze, Tommaso Schirinzi, Andrea Guerra, Alessandro Stefani, Mariangela Pierantozzi, Matteo Conti

## Abstract

**Objective:** To determine whether μ-phase–locked closed-loop repetitive transcranial magnetic stimulation (rTMS) induces distinct changes in corticospinal excitability and large-scale cortical functional connectivity (FC) compared with conventional open-loop stimulation.

**Methods:** Ten healthy volunteers underwent randomized, single-blind closed-loop, open-loop, and sham sessions in a crossover design. Closed-loop rTMS targeted the left primary motor cortex and was synchronized with the predicted negative peak of the individual μ rhythm; open-loop stimulation was delivered at 10 Hz. Each active session comprised 1,500 pulses. Motor-evoked potentials (MEPs) and resting-state high-density EEG were acquired before and after stimulation. Source-space weighted phase-lag index connectivity was analyzed across canonical frequency bands using the Network-Based Statistic.

**Results:** Both active protocols increased corticospinal excitability relative to baseline, whereas sham stimulation did not. The MEP increase was greater after closed-loop than open-loop stimulation (143.3±13.6% vs 132.5±16.0%; p=0.031). Open-loop rTMS reduced α-band FC. Closed-loop rTMS similarly reduced α-band FC but additionally increased β- and high-γ-band connectivity. Direct comparisons of individual FC changes confirmed significantly greater β- and high-γ increases after closed-loop than after open-loop stimulation, whereas α-band changes did not differ between the protocols. No significant changes in FC occurred after sham stimulation.

**Conclusions:** μ-phase–locked closed-loop rTMS enhanced corticospinal facilitation and induced broader, frequency-specific network reorganization than 10 Hz open-loop stimulation.

**Significance:** These preliminary findings indicate that stimulation timing relative to the ongoing cortical state may critically shape both local excitability and distributed network plasticity, supporting the further development of brain-state–dependent neuromodulation in future, larger, phase-controlled clinical studies.

## Introduction

Non-invasive brain stimulation (NIBS) has become a central tool in contemporary neuroscience research and clinical practice, enabling targeted, reversible modulation of cortical excitability. Techniques such as repetitive transcranial magnetic stimulation (rTMS) have been extensively applied to investigate brain function and to treat neurological and psychiatric disorders, including treatment-resistant depression, chronic pain, and motor deficits (Karabanov et al., 2016). Despite their widespread use, the clinical efficacy and reproducibility of NIBS protocols remain highly variable (Guerra et al., 2020a).

Traditionally, NIBS has been implemented according to an open-loop paradigm, in which stimulation parameters are predefined and delivered independently of the ongoing brain state. While this approach has enabled the development of standardized and reproducible protocols, it inherently neglects the dynamic nature of brain activity and the strong state-dependence of cortical excitability (Karabanov et al., 2016). As a consequence, identical stimulation protocols may lead to markedly different neurophysiological and behavioral outcomes across subjects, limiting both reliability and therapeutic impact.

Accumulating evidence indicates that the effects of TMS are critically shaped by the instantaneous neurophysiological state of the stimulated cortex. Variations in oscillatory activity, cortical excitability, and large-scale network configuration influence the neuronal response to external perturbations, thereby modulating the direction and magnitude of induced plasticity (Goldsworthy et al., 2021; Karabanov et al., 2021). In particular, oscillatory rhythms in the sensorimotor system have been shown to gate corticospinal excitability and responsiveness to stimulation, suggesting that the timing of TMS relative to endogenous brain rhythms represents a key determinant of its effectiveness (Guerra et al., 2020b, 2016). To overcome these limitations, a new paradigm of closed-loop stimulation has emerged in recent years. In this framework, stimulation is dynamically adapted in real time to the patient’s ongoing brain activity (Zrenner et al., 2016a). By tailoring stimulation to the current neurophysiological state, closed-loop approaches aim to increase clinical efficacy through more individualized interventions while simultaneously improving reproducibility and reliability by reducing the impact of inter-individual variability. However, current evidence is largely based on small-sample experimental studies and heterogeneous protocols, while well-controlled clinical trials are still needed to establish the true superiority and large-scale applicability of closed-loop stimulation (Karabanov et al., 2021; Zrenner et al., 2016a).

Beyond local changes in cortical excitability, NIBS-induced plasticity is increasingly understood as a network-level phenomenon (Makkinayeri et al., 2025). Stimulation of a single cortical target may propagate through structurally and functionally connected regions, leading to distributed changes in cortico-cortical communication. In this context, EEG-based functional connectivity (FC) provides a valuable approach to capture rapid stimulation-induced changes in large-scale brain networks with high temporal resolution. In particular, source-space EEG connectivity allows the investigation of frequency-specific interactions between cortical regions (Conti et al., 2026a, 2025b, 2025a, 2023), offering a direct window into the oscillatory mechanisms through which rTMS may modulate brain dynamics. This is especially relevant for closed-loop stimulation protocols based on endogenous rhythms, as their putative advantage lies in their ability to interact with ongoing oscillatory activity and reshape network synchronization in a state-dependent manner.

Based on this evidence, the present study aimed to characterize the effects of non-invasive closed-loop rTMS targeting the primary motor cortex (M1), phase-locked with the individual negative peak of the μ rhythm (8–13 Hz), on cortico-cortical EEG-based FC in healthy subjects. To determine whether these effects were specifically related to phase-dependent, state-dependent modulation of ongoing cortical oscillations, FC changes induced by closed-loop rTMS were directly compared with those observed after conventional open-loop 10-Hz rTMS. Sham stimulation was used as a further control condition.

## Materials and methods

### Subjects and study protocol

Ten young healthy volunteers were recruited at the University of Rome “Tor Vergata” (Rome, Italy). Inclusion criteria were: (i) age >18 years; and (ii) right-handedness. Exclusion criteria were: (i) history of neurological or systemic disorders; (ii) use of neuroactive drugs; and (iii) contraindications to TMS, including intracranial implanted devices, history of epilepsy or seizures, metallic implants in the head or neck region, or other standard safety contraindications.

All participants underwent high-density EEG (HD-EEG) recordings and rTMS delivered under three experimental conditions: i) closed-loop, ii) open-loop, and iii) sham. Neurophysiological assessments were performed before and after each stimulation condition. Resting-state HD-EEG and motor-evoked potentials (MEPs) were first acquired at baseline before rTMS (T0). Following stimulation, resting-state HD-EEG was recorded approximately 15 minutes after the end of the protocol, while corticospinal excitability was assessed approximately 20 minutes after stimulation, immediately after completion of the 5-minute post-stimulation EEG recording.

Each participant completed three experimental visits, consisting of a single rTMS session, with a one-week washout interval between visits. The order of the three stimulation conditions was randomized across participants. Participants were blinded to the stimulation condition, resulting in a single-blind experimental design. To ensure optimal reproducibility and minimize environmental variability, all experimental sessions were conducted under standardized conditions in a quiet and comfortable room during the morning hours.

The study was conducted in accordance with the Declaration of Helsinki and approved by the Ethics Committee of the Policlinico Tor Vergata (Protocol Code: RS 16/17). Written informed consent was obtained from all participants.

### HD-EEG acquisition and FC Analysis

HD-EEG was acquired for 5 minutes at a sampling rate of 1024 Hz using a 128-channel system (Brain Products GmbH, ActiCHamp Plus). Signals were band-pass filtered from 0.5-100 Hz and notch-filtered at 50 Hz. Electrodes were positioned according to the international 10–05 system(Oostenveld and Praamstra, 2001), and electrode impedance was maintained below 5 kΩ. Consistent with previous EEG studies(Yassine et al., 2023), recordings were performed during an eyes-closed (EC) resting-state (RS) condition: participants were asked to keep their eyes closed while remaining awake for 5 minutes.

For visualization purposes, EC data were segmented into 30-s epochs(Yassine et al., 2022), and the first epoch of each recording was discarded. To preserve temporal continuity and avoid artificial discontinuities that could bias phase-based connectivity measures, we manually selected the first six consecutive low-artifact epochs (total duration: 180 s) for subsequent analyses (Conti et al., 2026b).

Independent component analysis (ICA) was used to remove the residual EEG artifacts(Hyvärinen and Oja, 2000). Then, we proceeded with EEG source localization. EEG channels and MRI template (ICBM152) were co-registered through the identification of the same anatomical landmarks, and the Boundary Element Method (BEM)(Jatoi et al., 2016) was used to solve the forward problem. We used weighted minimum-norm estimation (wMNE) to solve the inverse problem (Grech et al., 2008). The sources obtained were divided into 68 brain regions, using the Desikan–Killiany atlas (Desikan et al., 2006).

FC was calculated in source space using weighted phase lag index (wPLI) values, a measure known to reduce conduction and noise artifacts and bias from small samples(Hardmeier et al., 2014). Phase information from the preprocessed signals had been computed using the Hilbert transform in δ (0.5-4 Hz), θ (4-8 Hz), α (8-13 Hz), β (13-30 Hz), low-γ (30-50 Hz), and high-γ (50-100 Hz) frequency bands. According to Welch’s method, dynamic FC matrices were computed for each pair of regions across the above EEG bands, using 1-second segments with 50% overlap. Of note, this method is not affected by the length of the displayed epochs (Jwo et al., 2021). Those matrices were averaged across time epochs to obtain static FC matrices.

### Transcranial Magnetic Stimulation Techniques

TMS was delivered over the left primary motor cortex using a figure-of-eight coil. MEPs were recorded from the right abductor pollicis brevis (APB) muscle using surface electrodes in a belly–tendon montage. The coil was positioned tangentially to the scalp over the APB motor hotspot, with the handle directed backward at approximately 45° from the midline. The optimal stimulation site was identified as the scalp position producing the largest and most consistent MEPs at the lowest stimulation intensity and was marked to ensure reproducible coil placement across sessions.

Resting and active motor thresholds were determined according to international guidelines (Rossini et al., 2015) and expressed as a percentage of the maximum stimulator output. Corticospinal excitability was assessed before stimulation (T0) and approximately 20 min after each stimulation condition (T1), following the post-rTMS resting-state EEG recording. At T0, the test stimulus intensity was adjusted to evoke a mean MEP amplitude of approximately 1 mV in the relaxed APB and was kept unchanged at T1. Ten MEPs were recorded at each time point using randomly jittered interstimulus intervals of 5–7 s. Mean peak-to-peak MEP amplitude was calculated separately at T0 and T1, and post-stimulation changes were expressed as a percentage of baseline. Further technical details are provided in the Supplementary Methods.

### rTMS protocol

In real (closed- and open-loop) conditions, stimulation consisted of 30 blocks of 50 pulses each, separated by 20-second pauses, for a total of 1500 pulses per session.

For the closed-loop condition, EEG activity was recorded in real time from the left (dominant) primary sensorimotor cortex. The active electrode was positioned over C3 according to the international 10–20 system, the reference electrode on the contralateral mastoid, and the ground electrode over Pz. EEG signals were acquired using a Digitimer D360 amplifier, digitized with a 16-bit AD converter (CED 1401, Cambridge Electronic Design, UK), and streamed in real time to the Simulink environment (MathWorks), where a custom-written closed-loop algorithm for oscillatory phase estimation and TMS triggering was implemented. At the beginning of each experimental session, a 60-second resting-state EEG recording was acquired in the absence of stimulation. Power spectral density was computed for this baseline segment to identify the subject-specific μ rhythm, defined as the dominant peak in the 8–13 Hz frequency range. This individual μ frequency was then used as the reference frequency for all subsequent steps of the closed-loop algorithm, where the system continuously monitored the EEG signal band-pass filtered around the subject-specific μ rhythm. The closed-loop algorithm was activated only when a stable oscillatory pattern was detected, defined as the presence of a regular and coherent μ oscillation for at least three consecutive cycles. This stability criterion was adopted to avoid stimulation during transient, noisy, or poorly representative oscillatory events. Once the stability criterion was fulfilled, the filtered EEG signal was processed using the Hilbert transform to obtain the analytic signal and estimate the instantaneous phase of the ongoing μ oscillation. Based on the current phase and the previously estimated individual μ frequency, the algorithm performed a forward phase prediction to estimate the timing of the subsequent negative peak of the oscillation, assuming a periodicity of 1/f, where f corresponded to the individual μ frequency. The first rTMS pulse of each stimulation block was synchronized with the predicted negative peak of the individual μ rhythm (Zrenner and Ziemann, 2024a). Stimulation continued for 50 pulses at the subject-specific μ frequency. This procedure was repeated 30 times, with a 20-second pause between blocks, for a total of 1500 pulses. The closed-loop pipeline was summarized in *Figure 1*.

**Figure 1.**
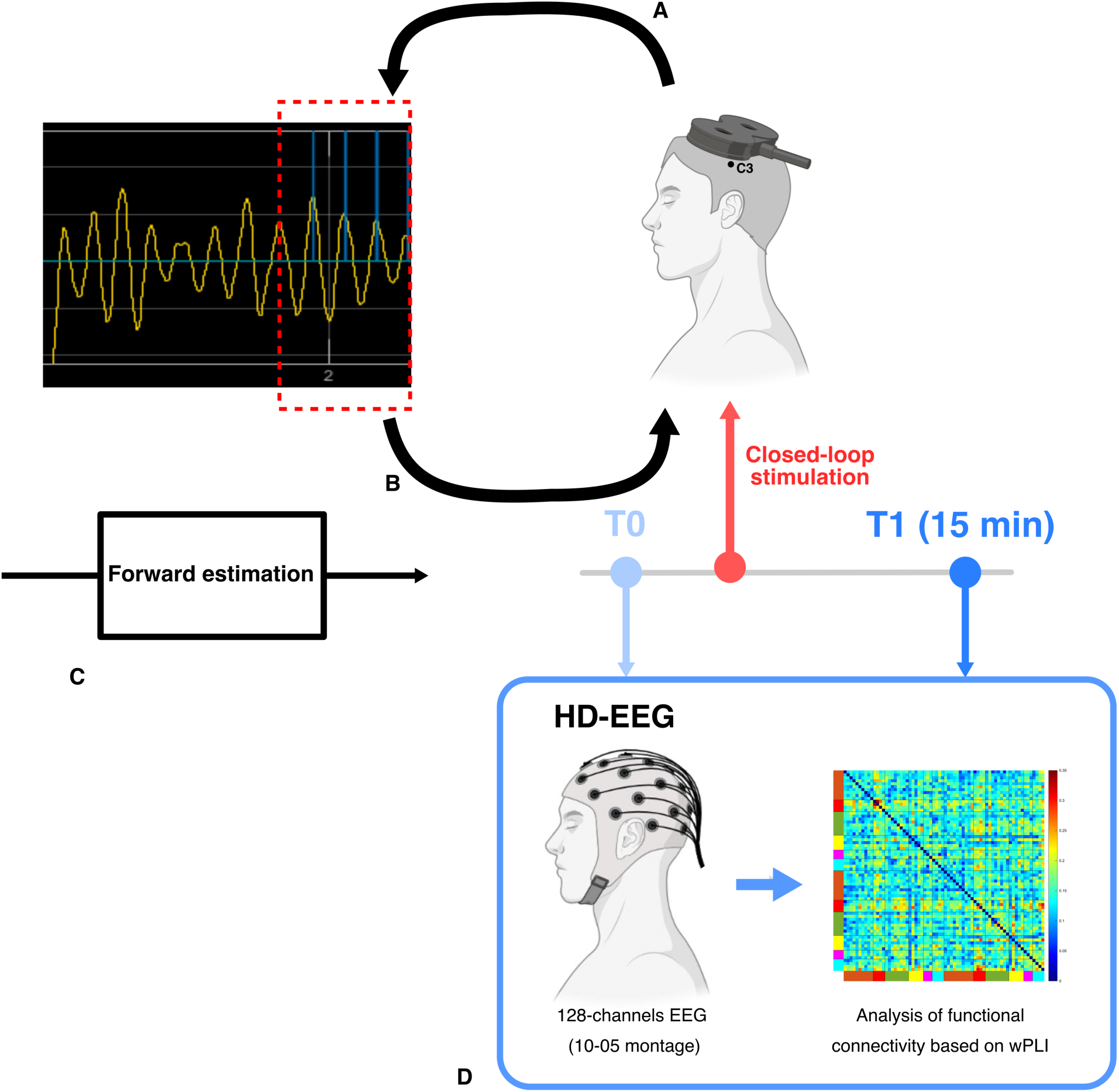
Experimental pipeline of closed-loop TMS and HD-EEG functional connectivity analysis. (A) A single-channel EEG signal was recorded over the left primary sensorimotor cortex, with the active electrode positioned over C3. The ongoing μ rhythm was monitored in real time to identify stable oscillatory activity. (B) After detection of a stable μ rhythm, a forward-phase-estimation algorithm was used to predict the timing of the upcoming negative peak, enabling phase-dependent triggering of TMS pulses. (C) Closed-loop stimulation was delivered by synchronizing TMS with the predicted phase of the endogenous μ rhythm. (D) HD-EEG recordings were acquired at baseline (T0) and 15 minutes after stimulation (T1). Source-space functional connectivity was computed using the weighted phase lag index (wPLI) to assess stimulation-induced changes in cortical network organization.

In the open-loop condition, rTMS was delivered using the same temporal structure and total number of pulses as the closed-loop condition, but without phase-dependent synchronization to the endogenous μ rhythm. Specifically, stimulation was delivered at a fixed frequency of 10 Hz, with 50 pulses per block for 30 blocks, separated by 20-second pauses, for a total of 1500 pulses. In this condition, pulse timing was therefore independent of the ongoing cortical oscillatory phase, while the number of pulses and the block structure were kept identical to those in the closed-loop protocol.

In the sham condition, the same experimental timeline and block structure were applied. Sham stimulation consisted of 30 blocks of 50 sham pulses separated by 20-second pauses. The figure-of-eight coil was positioned at 90° relative to the scalp, a sham procedure commonly adopted in rTMS studies to markedly reduce the induced cortical electric field while preserving the acoustic click associated with stimulation(Duecker and Sack, 2015). In addition, synchronized scalp electrical stimulation was delivered through two surface electrodes with each sham pulse to reproduce the somatosensory sensations associated with active stimulation. The sham condition matched the active conditions in terms of pulse number, session duration, acoustic stimulation, and somatosensory input, while minimizing effective cortical stimulation (Mennemeier et al., 2009).

### Statistical Analysis

Baseline motor thresholds and unconditioned MEP amplitudes were compared across closed-loop, open-loop, and sham conditions using separate one-way repeated-measures ANOVAs. Post-stimulation MEP amplitudes, recorded approximately 20 minutes after stimulation, were normalized to the corresponding baseline value as (post/pre)×100 and compared across conditions using a one-way repeated-measures ANOVA. Significant effects were followed by paired comparisons with Benjamini–Hochberg false discovery rate correction. Normalized MEP values were also compared with 100% using one-sample t-tests, with separate FDR correction. Greenhouse–Geisser correction was applied when sphericity was violated. Significance was set at an FDR-adjusted p<0.05.

FC changes between T0 and T1 were assessed separately for each stimulation condition and frequency band using the Network-Based Statistic (NBS) (Zalesky et al., 2010), with a primary cluster-forming threshold of t=2.9, 5,000 permutations, and an NBS-corrected significance level of p<0.05. Mean network connectivity (mNC) was calculated for significant components to describe the direction and magnitude of the effects. To compare stimulation-induced FC changes across conditions, individual-difference matrices were calculated as ΔFC = FC_T1_ − FC_T0_ and entered into paired NBS comparisons between closed-loop, open-loop, and sham stimulation. Both directional contrasts were evaluated when appropriate. Further methodological details are provided in the Supplementary Methods.

Statistical analyses were performed using MATLAB R2025b and the NBS toolbox. Graphs were generated using custom-written scripts in MATLAB R2023b and R.

No formal a priori sample-size calculation was performed. Given the protocol’s proof-of-concept nature and technical complexity, the sample size was primarily determined by feasibility. Nevertheless, the randomized crossover design adopted here, in which each participant underwent closed-loop, open-loop, and sham stimulation, was intended to reduce inter-individual variability and maximize sensitivity to condition-specific neurophysiological effects.

## Results

The study population had a mean age of 29.4 ± 3.1 years (range 25–35) and included 4 males and 6 females.

### Changes in corticospinal excitability following open-loop, closed-loop, and sham stimulation

Baseline unconditioned MEP amplitudes were comparable across stimulation conditions (closed-loop: 1.031 ± 0.115 mV; open-loop: 1.002 ± 0.077 mV; sham: 1.000 ± 0.077 mV), with no significant main effect of session (F=0.27, p=0.76). FDR-corrected pairwise comparisons likewise showed no significant baseline differences between closed-loop and open-loop stimulation (p=0.94), closed-loop and sham stimulation (p=0.94), or open-loop and sham stimulation (p=0.96).

MEP amplitudes measured approximately 20 minutes after stimulation, normalized to the corresponding baseline, differed significantly across stimulation conditions (F=18.98, p<0.001). This effect remained significant after Greenhouse–Geisser correction (p<0.001). Normalized MEP amplitudes increased to 143.3 ± 13.6% of baseline following closed-loop stimulation and to 132.5 ± 16.0% following open-loop stimulation, whereas no relevant change was observed following sham stimulation (97.4 ± 17.7%).

FDR-corrected post-hoc comparisons showed that normalized MEP amplitudes were significantly greater following closed-loop than open-loop stimulation (mean difference: 10.9, p=0.031). MEP amplitudes were also significantly greater following closed-loop than sham stimulation (mean difference: 45.9, p=0.001) and following open-loop than sham stimulation (mean difference: 35.0, p=0.008).

Compared with the reference value of 100%, indicating no change from baseline, MEP amplitudes were significantly increased following closed-loop stimulation (mean change: +43.3, p<0.001) and open-loop stimulation (mean change: +32.5, p<0.001). Conversely, MEP amplitudes following sham stimulation did not differ from baseline (mean change: −2.6, p=0.66). Differences in MEP amplitude were summarized in *Figure 2*.

**Figure 2.**
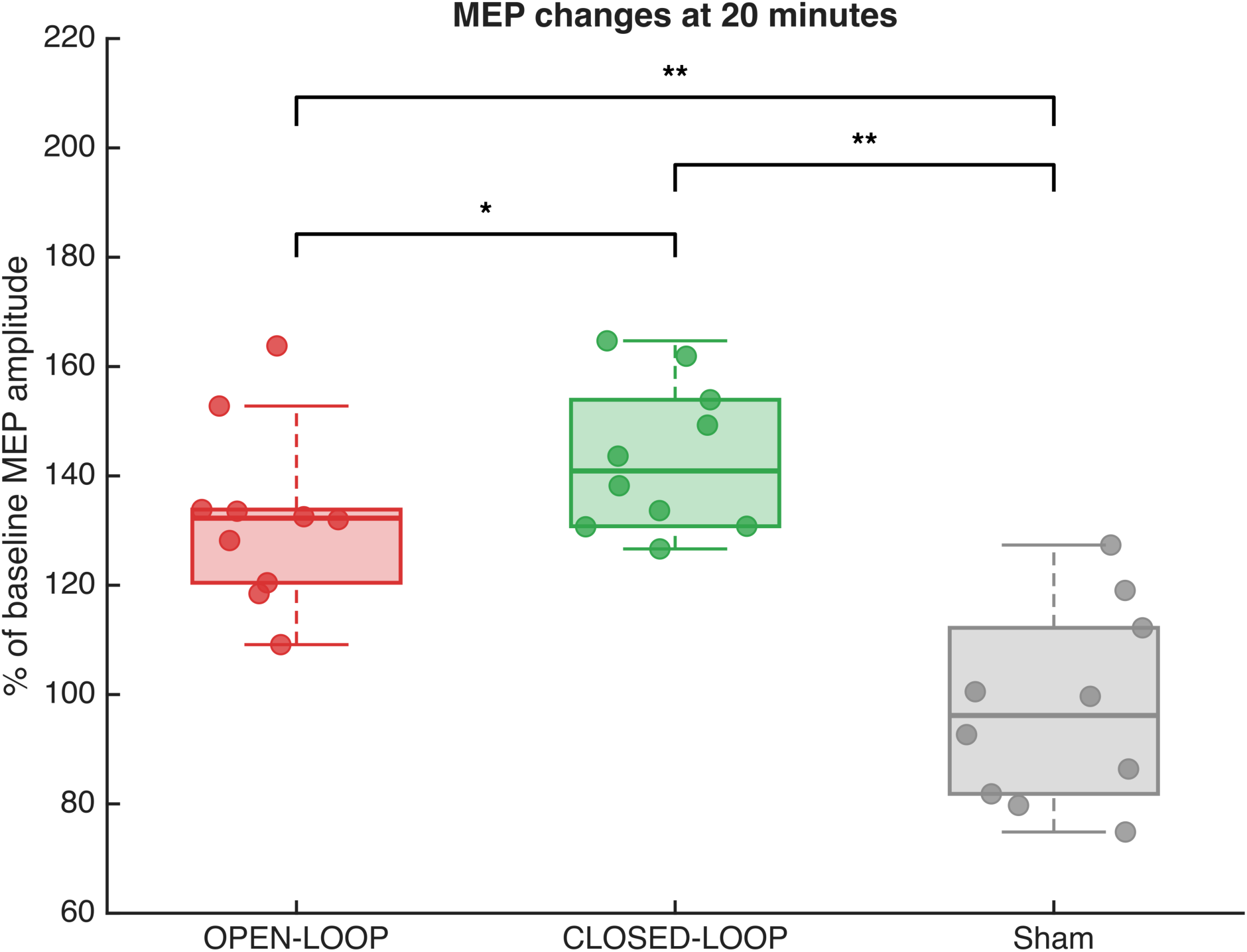
Changes in corticospinal excitability 20 minutes after open-loop, closed-loop, and sham stimulation. Motor-evoked potential (MEP) amplitudes recorded 20 minutes after stimulation are expressed as percentages of the corresponding baseline values. Individual dots represent single participants. Boxplots show the median and interquartile range, with whiskers extending to the most extreme values within 1.5 times the interquartile range. MEP amplitudes were significantly higher after both open-loop and closed-loop stimulation than after sham stimulation, and were also significantly higher after closed-loop than after open-loop stimulation. Brackets indicate significant pairwise comparisons. p < 0.05; p < 0.01. MEP = motor-evoked potential.

### Changes in EEG-based functional connectivity following open-loop, closed-loop, and sham stimulation

#### Open-loop stimulation

A significant NBS network was identified in the α band (t=2.9, p<0.001), where FC was reduced after the stimulation (T0>T1) (*Figure 3, Panels A-B*). This network comprised 59 nodes and 120 edges and showed a slight left-hemispheric predominance. At the nodal level, the most connected hub was the left lingual cortex (degree=15), followed by the right cuneus (degree=13), the left precentral gyrus (degree=11), and the right and left isthmus cingulate cortices (degree=9 and 8, respectively). Additional highly represented nodes included the left pericalcarine cortex, right postcentral gyrus, right paracentral lobule, right precuneus, and right superior parietal cortex (all degree=7), as well as the left temporal pole, left entorhinal cortex, left cuneus, right precentral gyrus, and right supramarginal gyrus (all degree=6). At the lobar level, the greatest contribution in terms of overall degree arose from occipital regions, followed by temporal, sensorimotor, fronto-insular, limbic, and parietal regions. In terms of connections, the largest proportion of significant links involved occipital-limbic connections (10.0%), followed by occipital-fronto-insular connections (9.2%), within-sensorimotor connections (8.3%), and temporal-fronto-insular connections (7.5%). Additional contributions were observed for occipital-sensorimotor, occipital-temporal, and limbic-temporal connections (6.7% each), as well as temporal-sensorimotor and parietal-temporal connections (5.8% each). Finally, the mNC of the identified component was also significantly lower at T1 than at T0 (t=23.33, p<0.001) (*Figure 3, Panel C*).

**Figure 3.**
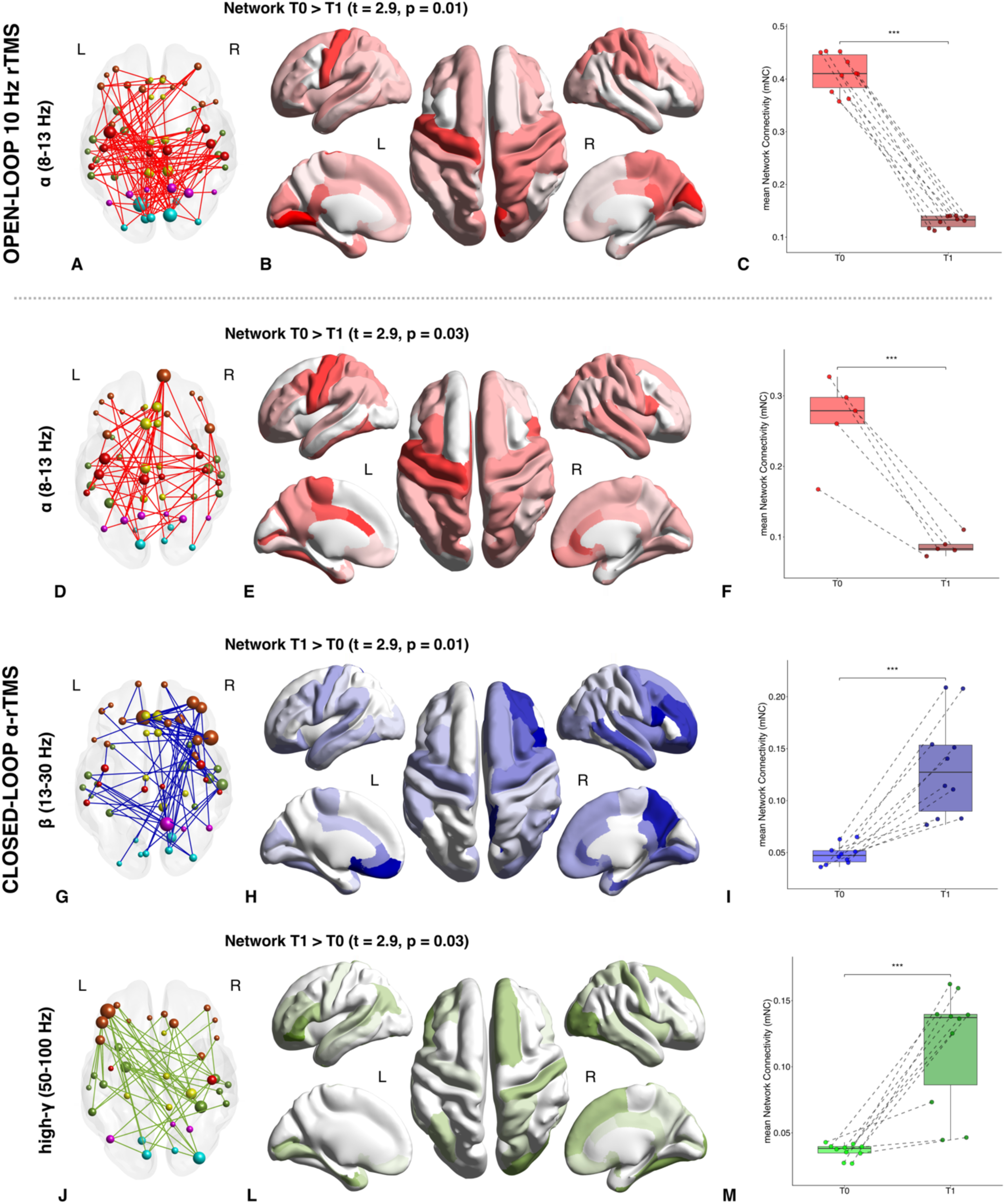
Frequency-specific functional connectivity changes induced by open-loop and closed-loop rTMS. (A–C) α-band (8–13 Hz) NBS network identified following open-loop 10-Hz rTMS, showing significantly greater functional connectivity at T0 than at T1 (T0 > T1; primary threshold t = 2.9, NBS-corrected p = 0.01). (D–F) α-band NBS network identified following closed-loop α-rTMS, also showing greater functional connectivity at T0 than at T1 (T0 > T1; primary threshold t = 2.9, NBS-corrected p = 0.03). (G–I) β-band (13–30 Hz) NBS network identified following closed-loop α-rTMS, showing greater functional connectivity at T1 than at T0 (T1 > T0; primary threshold t = 2.9, NBS-corrected p = 0.01). (J–M) High-γ-band (50–100 Hz) NBS network identified following closed-loop α-rTMS, showing greater functional connectivity at T1 than at T0 (T1 > T0; primary threshold t = 2.9, NBS-corrected p = 0.03). For each frequency band and stimulation condition, the left panel displays the significant network topology, with node size proportional to degree centrality, the central panel shows its cortical projection, and the right panel displays the paired distribution of mean network connectivity values at T0 and T1. Dashed lines connect measurements from the same participant. Asterisks indicate significant paired T0–T1 differences. L = left hemisphere; mNC = mean network connectivity; NBS = network-based statistic; R = right hemisphere; rTMS = repetitive transcranial magnetic stimulation; T0 = pre-stimulation assessment; T1 = post-stimulation assessment.

No significant differences were observed in the other frequency bands.

#### Closed-loop stimulation

A significant NBS network was identified in the α band (t=2.9, p=0.028), where FC was reduced after stimulation (T0>T1) (*Figure 3, Panels D-E*). This network comprised 51 nodes and 71 edges and showed a slight left-hemispheric predominance. At the nodal level, the most connected hub was the right frontal pole (degree=8), followed by the left precentral gyrus and left caudal anterior cingulate cortex (degree=6 each). Additional highly represented nodes included the left postcentral gyrus, left paracentral lobule, left fusiform gyrus, left posterior cingulate cortex, left pericalcarine cortex, right pars opercularis, right transverse temporal cortex, and right rostral anterior cingulate cortex. At the lobar level, the greatest contribution in terms of overall degree arose from the frontal, sensorimotor, temporal, and limbic regions. In terms of connections, the largest proportion of significant links involved limbic-temporal connections (14.1%), followed by within-sensorimotor connections and temporal-frontal connections (11.3%), and limbic-frontal connections (8.5%). Additional contributions were observed for within-frontal, parietal-frontal, and occipital-sensorimotor connections (5.6%), while parietal-sensorimotor, within-parietal, and within-occipital links were not represented. Finally, the mNC of the identified component was also significantly lower at T1 than at T0 (t=6.03, p<0.001) (*Figure 3, Panel F*).

Moreover, a significant NBS network was observed in the β band (t=2.9, p=0.015), showing greater FC after stimulation (T1>T0) (*Figure 3, Panels G-H*). This network comprised 53 nodes and 82 edges, with right-hemispheric predominance in the nodal degree distribution. The most connected hubs were the left medial orbitofrontal cortex and the right pars opercularis (degree=9 each), followed by the right precuneus (degree=8) and the right rostral middle frontal gyrus (degree=7). Additional highly represented nodes included the right pars orbitalis, right entorhinal cortex, and right middle temporal gyrus (degree=6 each), as well as the right lateral orbitofrontal cortex, right caudal middle frontal gyrus, right transverse temporal cortex, right rostral anterior cingulate cortex, and right lingual gyrus (degree=5 each). At the lobar level, the greatest contribution in terms of overall degree arose from frontal regions, followed by temporal regions and limbic regions. Consistent with the right-sided predominance, right-hemispheric nodes accounted for 54.7% of active nodes but 67.8% of the total degree. In terms of connections, the largest proportion of significant links involved within-frontal connections (22.0%), followed by limbic–frontal connections (14.6%). Additional relevant contributions were observed for parietal–temporal, occipital–frontal, and occipital–temporal connections (8.5% each), as well as within-temporal connections (7.3%) and temporal–frontal connections (6.1%). Conversely, within-limbic, parietal–sensorimotor, within-parietal, occipital–parietal, and within-occipital connections were not represented. Finally, the mNC of the identified component was also significantly higher at T1 than at T0 (t = −5.39, p < 0.001) (*Figure 3, Panel I*).

Finally, we found an NBS network in the high-γ band (t=2.9, p=0.03), where FC was significantly increased after stimulation (T1>T0) (*Figure 3, Panels J-L*). This network comprised 37 nodes and 53 edges and showed a relatively balanced hemispheric distribution. At the nodal level, the most connected hub was the left pars orbitalis (degree=8), followed by the left pars triangularis, right fusiform gyrus, and right lateral occipital cortex (degree=6 each). Additional highly represented nodes included the left entorhinal cortex and right parahippocampal gyrus (degree=5 each), as well as the left pars opercularis, left pericalcarine cortex, right superior frontal gyrus, right postcentral gyrus, and right transverse temporal cortex (degree=4 each). At the lobar level, the greatest contribution in terms of overall degree arose from frontal regions, followed by temporo-parietal and occipital regions. Hemispheric involvement was relatively balanced (left-hemispheric 49.1% of nodes). In terms of edges, the largest proportion of significant links involved temporal–frontal connections (28.3%), followed by limbic–frontal connections (18.9%). Additional relevant contributions were observed in occipital–frontal and within-temporal connections (9.4% each) and in fronto–parietal connections (7.5%). Furthermore, the mNC of the identified component was also significantly higher at T1 than at T0 (t = −5.64, p < 0.001) (*Figure 3, Panel M*).

No significant differences were observed in the other frequency bands.

#### Sham stimulation

No significant differences were found between T0 and T1 in any frequency bands.

#### Direct comparison of stimulation-induced FC changes across conditions

Direct comparison of the individual ΔFC matrices revealed significant differences between closed-loop and open-loop stimulation in both the β and high-γ bands. In the β band, the stimulation-induced increase in FC was greater following closed-loop than open-loop stimulation (ΔFC_closed-loop_>ΔFC_open-loop_; t=2.9, p=0.005). The network comprised 64 nodes and 160 edges. Its regional distribution largely overlapped with that observed in the closed-loop T1>T0 analysis, with predominant involvement of frontal, temporal, and limbic regions and substantial long-range fronto-temporal and fronto-limbic connectivity. A similar pattern emerged in the high-γ band, where the FC increase was also significantly greater following closed-loop than open-loop stimulation (ΔFC_closed-loop_>ΔFC_open-loop_; t=2.9, p=0.013). This component included 59 nodes and 104 edges. The anatomical distribution was broadly consistent with the high-γ network identified in the closed-loop T1>T0 comparison, prominently involving frontal and temporal regions, with additional contributions from parietal and occipital areas and widespread long-range connections. No significant difference between closed-loop and open-loop stimulation was detected in the α/μ band.

Compared with sham stimulation, open-loop rTMS produced a significantly greater reduction in α/μ-band connectivity, whereas closed-loop rTMS produced significantly greater changes in α/μ, β, and high-γ FC. The direction of these effects was consistent with the corresponding pre–post analyses: greater α/μ-band connectivity reduction after both active conditions and greater β- and high-γ-band connectivity increases after closed-loop stimulation. No significant between-condition differences were detected in the remaining frequency bands.

## Discussion

In the present study, we investigated the effects of conventional open-loop 10-Hz rTMS and μ-phase–locked closed-loop rTMS applied over the primary motor cortex on corticospinal excitability and large-scale EEG-based FC in healthy subjects. Both active stimulation conditions produced significant neurophysiological effects, whereas sham stimulation did not. Open-loop stimulation increased corticospinal excitability and reduced α-band FC within a widespread network involving sensorimotor, frontal, and temporo-parietal regions. Closed-loop stimulation produced a larger increase in corticospinal excitability and was associated with a reduction in α-band FC in a similar network, accompanied by additional increases in β- and high-γ-band FC. In particular, the β-band network predominantly involved frontal, temporal, and limbic regions, whereas the high-γ-band network mainly included frontal and temporo-parietal areas. Importantly, direct comparisons of stimulation-induced changes confirmed that the β- and high-γ increases were significantly greater after closed-loop than open-loop stimulation, whereas the α-band reduction did not differ between the two active protocols. These findings indicate that rhythmic stimulation itself may account for the shared α modulation, while phase-dependent timing selectively enhances corticospinal facilitation and higher-frequency network reorganization.

Open-loop rTMS produced a widespread reduction in α-band connectivity involving occipital, temporal, sensorimotor, fronto-insular, limbic, and parietal regions. This finding is consistent with the capacity of rhythmic stimulation delivered within the α range to perturb the ongoing synchronization of α networks. Sensorimotor α/μ activity has traditionally been interpreted as an inhibitory or idling rhythm, whose attenuation accompanies motor preparation, movement, motor imagery, and increased cortical engagement (Pfurtscheller et al., 1996). Accordingly, the post-stimulation reduction in α/μ connectivity may reflect a persistent desynchronization of the stimulated sensorimotor system and its connected cortical regions. However, the absence of significant β- or high-γ changes after open-loop stimulation suggests that frequency-matched stimulation delivered independently of the ongoing oscillatory phase mainly affected the stimulated frequency range, without producing a comparably broad cross-frequency network reorganization.

Closed-loop stimulation reproduced the reduction in α/μ-band connectivity but additionally increased connectivity in the β and high-γ bands. The lack of a significant difference between closed-loop and open-loop stimulation in the direct α/μ comparison indicates that phase-locking did not further amplify the α reduction itself. Thus, the α/μ effect may primarily reflect the common rhythmic characteristics of the two active protocols, including stimulation frequency, pulse number, and temporal structure. In contrast, the selective emergence of β- and high-γ activity after closed-loop stimulation suggests that aligning stimulation with the ongoing μ phase recruited additional network mechanisms beyond those induced by conventional 10-Hz rTMS.

This interpretation is strengthened by the direct comparisons of the individual ΔFC matrices. In the β band, closed-loop stimulation produced a significantly greater increase in FC than open-loop stimulation across a widespread network, involving frontal, temporal, and limbic regions. The anatomical distribution broadly overlapped with the β-band network identified in the closed-loop pre–post analysis. β oscillations are implicated in sensorimotor integration, top-down control, maintenance of the current motor state, and long-range cortical communication (Engel and Fries, 2010; Spitzer and Haegens, 2017). The greater β-band effect after phase-locked stimulation may therefore reflect enhanced integration of the post-stimulation motor network state rather than a simple modulation of the stimulated μ rhythm. A similar pattern was observed in the high-γ band. Closed-loop stimulation induced a significantly greater FC increase than open-loop stimulation within a network prominently involving frontal and temporo-parietal cortices. This distribution was also broadly consistent with the high-γ network observed in the closed-loop T1>T0 analysis. High-γ activity is commonly associated with fast cortical processing, local neuronal engagement, and efficient functional communication (Fries, 2009; Ray and Maunsell, 2015). The present results suggest that phase-dependent stimulation may facilitate faster modes of cortical interaction that are not substantially recruited by frequency-matched open-loop stimulation.

The corticospinal excitability findings provide an independent neurophysiological outcome supporting the effectiveness of both active stimulation protocols. MEP amplitudes increased significantly after both open-loop and closed-loop stimulation, whereas no relevant change was observed after sham stimulation. Moreover, the MEP increase was significantly greater following closed-loop than open-loop stimulation. This result is particularly relevant because stimulation was delivered at the negative peak of the individual μ rhythm, a phase previously associated with a state of relatively high corticospinal excitability compared with the opposite phase (Schaworonkow et al., 2019; Zrenner et al., 2016b). Repeated stimulation during this excitability window may have increased the probability of inducing facilitatory plasticity, thereby producing a larger corticospinal response than phase-independent stimulation.

Taken together, the MEP and FC findings support a brain-state–dependent interpretation of the closed-loop effect. Open-loop stimulation was sufficient to increase corticospinal excitability and perturb α synchronization. However, closed-loop stimulation enhanced the corticospinal response and produced additional β- and high-γ network changes that were significantly greater than those observed after open-loop stimulation. The convergence of these local and network-level outcomes suggests that phase-dependent stimulation may exploit transient windows of enhanced responsiveness to reinforce stimulation-induced plasticity and promote a broader reconfiguration of cortical communication. To note, the present findings should not be interpreted as evidence of simple oscillatory entrainment. If closed-loop stimulation had primarily strengthened the targeted μ oscillation, an increase rather than a reduction in α/μ connectivity might have been expected. Instead, both active protocols reduced α/μ connectivity, while closed-loop stimulation selectively increased higher-frequency FC. A more plausible interpretation is therefore that the μ rhythm acted as an online marker of cortical state rather than merely as a rhythm to be reinforced. By triggering stimulation during a negative μ phase associated with increased corticospinal responsiveness, the closed-loop protocol may have delivered repeated perturbations during a more plastic network configuration. This may have destabilized the prevailing α/μ synchronization while facilitating subsequent β- and high-γ network integration.

These results are in line with contemporary models of closed-loop neuromodulation, according to which stimulation efficacy depends not only on conventional parameters such as intensity and frequency, but also on the instantaneous physiological state of the stimulated cortex (Zrenner and Ziemann, 2024b). In this framework, the advantage of closed-loop stimulation arises from reducing the variability of the prestimulus brain state and repeatedly targeting a neurophysiologically favorable window. The present study extends this concept beyond single-pulse MEP modulation by showing that phase-locked stimulation can influence both corticospinal excitability and distributed cortico-cortical communication.

### Limitations and strengths

This study has several limitations. First, the sample size was small, and no formal a priori sample-size calculation was performed. The study should therefore be regarded as exploratory, and the observed effects require confirmation in larger cohorts. Second, closed-loop stimulation was tested only at the negative peak of the μ rhythm. The inclusion of stimulation at the positive peak or of both opposite phases within the same experimental design would have provided a stronger test of phase specificity. Third, EEG-based high-γ connectivity should be interpreted cautiously, as scalp-recorded high-frequency activity may be influenced by residual muscle or non-neural artifacts despite careful preprocessing, source reconstruction, and the use of wPLI. Finally, only short-term effects were assessed approximately 15–20 minutes after stimulation; therefore, the persistence and behavioral relevance of these changes remain unknown.

Despite these limitations, the study has several strengths. The randomized crossover design allowed each participant to serve as their own control, reducing inter-individual variability. The inclusion of both open-loop and sham conditions enabled us to distinguish the effects of rhythmic stimulation from those related to phase-dependent triggering, from nonspecific auditory and somatosensory effects, and from expectancy and repeated-measurement effects. The active protocols were matched for pulse number and temporal structure, allowing a direct assessment of the contribution of stimulation timing. Furthermore, the combined assessment of MEPs and HD-EEG source-space connectivity provided complementary local and network-level measures of stimulation-induced plasticity.

## Conclusions

Both open-loop and closed-loop rTMS increased corticospinal excitability and reduced α/μ-band FC. However, closed-loop stimulation produced significantly greater MEP facilitation and selectively induced greater increases in β- and high-γ connectivity than open-loop stimulation. These findings suggest that stimulation delivered during a negative μ-phase excitability window engages additional plasticity mechanisms beyond those produced by frequency-matched, phase-independent rTMS. Phase-dependent timing may therefore represent an important determinant of both local corticospinal responsiveness and large-scale network reorganization. Nevertheless, because the sample was small, these findings should be considered preliminary and require confirmation using larger phase-controlled designs.

## Acknowledgements

The authors would like to thank all the healthy volunteers who participated in the experimental sessions.

## Author Contribution

Federico Carparelli: Conceptualization, Methodology, Investigation, Writing - Original Draft; Clara Simonetta: Methodology, Investigation; Davide Mascioli: Investigation; Valerio Ferrari: Investigation; Silvio Bagetta: Investigation; Carlo Conti: Methodology, Software; Diego Centonze: Supervision; Tommaso Schirinzi: Writing - Review & Editing, Funding; Andrea Guerra: Methodology, Writing - Review & Editing; Alessandro Stefani: Supervision, Funding; Mariangela Pierantozzi: Writing - Review & Editing, Funding; Matteo Conti: Conceptualization, Methodology, Software, Formal analysis, Investigation, Writing - Original Draft.

## Data Availability

The raw data supporting the conclusions of this article will be made available by the corresponding author upon reasonable request.

## References

Conti M, Carparelli F, Bovenzi R, Ferrari V, Di Gioia B, Mercuri NB, et al. Exercise-induced changes in high-γ cortical functional connectivity and short-interval intracortical inhibition. Clinical Neurophysiology 2025a;173. 10.1016/j.clinph.2025.02.274.

Conti M, D’Onofrio V, Bovenzi R, Ferrari V, Di Giuliano F, Cerroni R, et al. Cortical Functional Connectivity Changes in the Body-First and Brain-First Subtypes of Parkinson’s Disease. Movement Disorders 2025b;40:254–65. 10.1002/mds.30071.

Conti M, Guerra A, Pierantozzi M, Bovenzi R, D’Onofrio V, Simonetta C, et al. Band-Specific Altered Cortical Connectivity in Early Parkinson’s Disease and its Clinical Correlates. Movement Disorders 2023;38:2197–208. 10.1002/mds.29615.

Conti M, Mascioli D, Simonetta C, Ferrari V, Bissacco J, Bagetta S, et al. Clinical, Biological, and Functional Connectivity Profile of Patients With De Novo Parkinson Disease Who Are *APOE* ε4 Carriers. Neurology 2026a;106. 10.1212/WNL.0000000000214449.

Conti M, Mascioli D, Simonetta C, Ferrari V, Bissacco J, Bagetta S, et al. Clinical, Biological, and Functional Connectivity Profile of Patients With De Novo Parkinson Disease Who Are *APOE* ε4 Carriers. Neurology 2026b;106. 10.1212/WNL.0000000000214449.

Desikan RS, Ségonne F, Fischl B, Quinn BT, Dickerson BC, Blacker D, et al. An automated labeling system for subdividing the human cerebral cortex on MRI scans into gyral based regions of interest. Neuroimage 2006;31. 10.1016/j.neuroimage.2006.01.021.

Duecker F, Sack AT. Rethinking the role of sham TMS. Front Psychol 2015;6. 10.3389/fpsyg.2015.00210.

Engel AK, Fries P. Beta-band oscillations-signalling the status quo? Curr Opin Neurobiol 2010;20. 10.1016/j.conb.2010.02.015.

Fries P. Neuronal gamma-band synchronization as a fundamental process in cortical computation. Annu Rev Neurosci 2009;32. 10.1146/annurev.neuro.051508.135603.

Goldsworthy MR, Hordacre B, Rothwell JC, Ridding MC. Effects of rTMS on the brain: is there value in variability? Cortex 2021;139. 10.1016/j.cortex.2021.02.024.

Grech R, Cassar T, Muscat J, Camilleri KP, Fabri SG, Zervakis M, et al. Review on solving the inverse problem in EEG source analysis. J Neuroeng Rehabil 2008;5:1–33. 10.1186/1743-0003-5-25.

Guerra A, López-Alonso V, Cheeran B, Suppa A. Variability in non-invasive brain stimulation studies: Reasons and results. Neurosci Lett 2020a;719:133330. 10.1016/j.neulet.2017.12.058.

Guerra A, López-Alonso V, Cheeran B, Suppa A. Solutions for managing variability in non-invasive brain stimulation studies. Neurosci Lett 2020b;719. 10.1016/j.neulet.2017.12.060.

Guerra A, Pogosyan A, Nowak M, Tan H, Ferreri F, Di Lazzaro V, et al. Phase Dependency of the Human Primary Motor Cortex and Cholinergic Inhibition Cancelation during Beta tACS. Cerebral Cortex 2016;26. 10.1093/cercor/bhw245.

Hardmeier M, Hatz F, Bousleiman H, Schindler C, Stam CJ, Fuhr P. Reproducibility of functional connectivity and graph measures based on the phase lag index (PLI) and weighted phase lag index (wPLI) derived from high resolution EEG. PLoS One 2014;9. 10.1371/journal.pone.0108648.

Hyvärinen A, Oja E. Independent component analysis: Algorithms and applications. Neural Networks 2000;13. 10.1016/S0893-6080(00)00026-5.

Jatoi MA, Kamel N, Faye I, Malik AS, Bornot JM, Begum T. BEM based solution of forward problem for brain source estimation. IEEE 2015 International Conference on Signal and Image Processing Applications, ICSIPA 2015 - Proceedings 2016:180–5. 10.1109/ICSIPA.2015.7412186.

Jwo DJ, Chang WY, Wu IH. Windowing Techniques, the Welch Method for Improvement of Power Spectrum Estimation. Computers, Materials and Continua 2021;67. 10.32604/cmc.2021.014752.

Karabanov A, Thielscher A, Siebner HR. Transcranial brain stimulation: closing the loop between brain and stimulation. Curr Opin Neurol 2016;29:397–404. 10.1097/WCO.0000000000000342.

Karabanov AN, Shindo K, Shindo Y, Raffin E, Siebner HR. Multimodal Assessment of Precentral Anodal TDCS: Individual Rise in Supplementary Motor Activity Scales With Increase in Corticospinal Excitability. Front Hum Neurosci 2021;15. 10.3389/fnhum.2021.639274.

Makkinayeri S, Guidotti R, Basti A, Woolrich MW, Gohil C, Pettorruso M, et al. Investigating brain network dynamics in state-dependent stimulation: A concurrent electroencephalography and transcranial magnetic stimulation study using hidden Markov models. Brain Stimul 2025;18. 10.1016/j.brs.2025.03.020.

Mennemeier MS, Triggs WJ, Chelette KC, Woods AJ, Kimbrell TA, Dornhoffer JL. Sham transcranial magnetic stimulation using electrical stimulation of the scalp. Brain Stimul 2009;2:168–73. 10.1016/j.brs.2009.02.002.

Oostenveld R, Praamstra P. The five percent electrode system for high-resolution EEG and ERP measurements. Clinical Neurophysiology 2001;112:713–9. 10.1016/S1388-2457(00)00527-7.

Pfurtscheller G, Stancák A, Neuper C. Event-related synchronization (ERS) in the alpha band - An electrophysiological correlate of cortical idling: A review. International Journal of Psychophysiology 1996;24. 10.1016/S0167-8760(96)00066-9.

Ray S, Maunsell JHR. Do gamma oscillations play a role in cerebral cortex? Trends Cogn Sci 2015;19. 10.1016/j.tics.2014.12.002.

Rossini PM, Burke D, Chen R, Cohen LG, Daskalakis Z, Di Iorio R, et al. Non-invasive electrical and magnetic stimulation of the brain, spinal cord, roots and peripheral nerves: Basic principles and procedures for routine clinical and research application: An updated report from an I.F.C.N. Committee. Clinical Neurophysiology 2015;126. 10.1016/j.clinph.2015.02.001.

Schaworonkow N, Triesch J, Ziemann U, Zrenner C. EEG-triggered TMS reveals stronger brain state-dependent modulation of motor evoked potentials at weaker stimulation intensities. Brain Stimul 2019;12. 10.1016/j.brs.2018.09.009.

Spitzer B, Haegens S. Beyond the status quo: A role for beta oscillations in endogenous content (RE)activation. ENeuro 2017;4. 10.1523/ENEURO.0170-17.2017.

Yassine S, Gschwandtner U, Auffret M, Achard S, Verin M, Fuhr P, et al. Functional Brain Dysconnectivity in Parkinson’s Disease: A 5-Year Longitudinal Study. Movement Disorders 2022;37:1444–53. 10.1002/mds.29026.

Yassine S, Gschwandtner U, Auffret M, Duprez J, Verin M, Fuhr P, et al. Identification of Parkinson’s Disease Subtypes from Resting-State Electroencephalography. Movement Disorders 2023;38:1451–60. 10.1002/mds.29451.

Zalesky A, Fornito A, Bullmore ET. Network-based statistic: Identifying differences in brain networks. Neuroimage 2010;53. 10.1016/j.neuroimage.2010.06.041.

Zrenner C, Belardinelli P, Müller-Dahlhaus F, Ziemann U. Closed-Loop Neuroscience and Non-Invasive Brain Stimulation: A Tale of Two Loops. Front Cell Neurosci 2016a;10. 10.3389/fncel.2016.00092.

Zrenner C, Belardinelli P, Müller-Dahlhaus F, Ziemann U. Closed-loop neuroscience and non-invasive brain stimulation: A tale of two loops. Front Cell Neurosci 2016b;10. 10.3389/fncel.2016.00092.

Zrenner C, Ziemann U. Closed-Loop Brain Stimulation. Biol Psychiatry 2024a;95:545–52. 10.1016/j.biopsych.2023.09.014.

Zrenner C, Ziemann U. Closed-Loop Brain Stimulation. Biol Psychiatry 2024b;95. 10.1016/j.biopsych.2023.09.014.

