## Supplementary Methods for "Closed-Loop rTMS Induces Frequency-Specific Cortical Network Reorganization Distinct from Open-Loop Stimulation in Healthy Humans"

**Transcranial Magnetic stimulation techniques**

TMS was performed with an 8-shaped coil (each loop diameter 7 cm; peak magnetic field 2.2 Tesla) connected with two Magstim magnetic Stimulators (Magstim Company, UK). MEPs to TMS were recorded from the abductor pollicis brevis (APB) muscle via surface electrodes applied in a belly-tendon bipolar montage. The left hemisphere was selected for stimulation, and the coil was placed at 7 cm lateral and 2 cm forward to Cz along the interlobe line, over the scalp region corresponding to the primary hand motor area contralateral to the target muscle. The coil was held tangentially to the scalp, with the handle pointing backward and 45° angle from the midline. The cortical site where MEPs with the lowest intensity (threshold) were consistently elicited in the contralateral APB was carefully located and marked with a dermographic pencil to maintain the optimal stimulating position throughout all TMS sessions.

In agreement with international standards(Rossini et al., 2015), the resting motor threshold (RMT) was defined as the lowest stimulus intensity that elicited a clear MEP of 100 μV in 50% of 10 consecutive stimuli at rest. The active motor threshold (AMT) was defined as the lowest stimulus intensity able to produce a MEP of 100 μV during tonic activation at about 20% of maximum voluntary contraction, across 50% of 10 consecutive stimuli. Both the RMT and the AMT were expressed as a percentage of the magnetic stimulator's maximal output (100%).

Corticospinal excitability was assessed before stimulation (T0) and approximately 20 minutes after the end of each stimulation condition (T1). The post-stimulation MEP assessment was performed after the 5-minute resting-state EEG recording acquired immediately following rTMS. At baseline, the test stimulus intensity was individually adjusted to evoke a mean MEP peak-to-peak amplitude of approximately 1 mV in the relaxed right APB muscle. The same stimulus intensity was maintained for the post-stimulation assessment. Ten consecutive MEPs were recorded with an interstimulus interval of [5–7 s, randomly jittered], while participants were instructed to remain completely relaxed. The mean MEP amplitude was calculated separately at T0 and T1 for each experimental condition. Stimulation-induced changes in corticospinal excitability were expressed as the percentage of the baseline MEP amplitude.

EMG activity was recorded from the right APB using surface electrodes taped in a belly-tendon montage. EMG signals were amplified and filtered (20 Hz–1 kHz) using Digitimer D360. EMG signals were recorded and stored on a laboratory PC (sampling rate of 5 kHz) using an analog-to-digital converter (ADC) AD1401 Plus (Cambridge Electronic Design). Off-line analyses were performed using dedicated software (Signal® version 4.00, Cambridge Electronic Design). The MEP peak-to-peak amplitude was measured within a 20–40 ms time window after the TMS artifact. Traces with background EMG activity exceeding 100 mV in the 200 ms time window preceding the TMS artifact were rejected online.

**Statistical Analysis**

Baseline motor thresholds and unconditioned MEP amplitudes were compared across stimulation conditions using separate one-way repeated-measures analyses of variance (ANOVAs), with session (closed-loop, open-loop, and sham) as the within-subject factor.

To evaluate stimulation-induced changes in corticospinal excitability, mean MEP amplitudes obtained approximately 20 minutes after stimulation were normalized to the corresponding pre-stimulation value for each participant and session according to the following formula: (post-stimulation MEP/pre-stimulation MEP)×100. Normalized post-stimulation MEP amplitudes were compared using a one-way repeated-measures ANOVA with session (closed-loop, open-loop, and sham) as the within-subject factor. Significant main effects were followed by paired post-hoc comparisons, with p-values adjusted using the Benjamini–Hochberg false discovery rate procedure.

As a complementary analysis, normalized MEP amplitudes in each stimulation condition were compared with the reference value of 100% (indicating no change from baseline) using one-sample t-tests. The resulting p-values were corrected separately using the Benjamini–Hochberg false discovery rate procedure. The Greenhouse–Geisser correction was applied when the assumption of sphericity was violated, as indicated by Mauchly's test. Statistical significance was set at an FDR-adjusted $p < 0.05$. Values are presented as mean ± SD.

Differences in cortico-cortical FC between T0 and T1 were assessed separately for each stimulation condition and for each frequency band using the Network-Based Statistic (NBS). NBS is a cluster-based statistical method used in several previous studies(Conti et al., 2024, 2023; De Micco et al., 2018; Yassine et al., 2022), which provides greater statistical power than standard univariate tests and traditional correction methods(Zalesky et al., 2010).

Specifically, in this study, for each frequency band and stimulation condition, we performed an edge-wise paired t-test to compute the network size whose edges had greater weights than the defined initial t threshold. We then performed a permutation test, randomly assigning all subjects to one of the three groups, maintaining each group size N-1 times, and computed the maximum sizes of networks whose edges had weights greater than the threshold, resulting in an empirical null distribution of maximum sizes. Then, we assigned the p-value of the network to the fraction of occurrences whose sizes were larger than the network size in the original assignment. In this study, we used $t = 2.9$ (lower t with $p<0.01$ at the given degrees of freedom) and 5000 as the initial threshold and the number of permutations (N), respectively. The threshold was incremented by 0.1 steps. If no significant component was detected at $t = 2.9$, the cluster-forming threshold was progressively increased by 0.1 to determine whether a more restricted suprathreshold component survived NBS correction. This procedure was continued until no connected component was identified. Network components were considered statistically significant only when the permutation-based NBS-corrected p-value was $< 0.05$. For each significant NBS component, mean network connectivity (mNC) was calculated as the average FC value across all edges included in the identified network. The mNC was then extracted at T0 and T1 and compared using paired t-tests to confirm the direction and magnitude of the stimulation-induced effect within each condition.

Moreover, to directly compare the magnitude and direction of stimulation-induced FC changes across conditions, individual difference matrices were subsequently calculated for each subject, frequency band, and stimulation condition as $\Delta FC = FC_{T1} - FC_{T0}$. These ΔFC matrices were then entered into paired NBS comparisons between closed-loop and open-loop stimulation, closed-loop and sham stimulation, and open-loop and sham stimulation. For each pairwise comparison, edge-wise paired t-tests were performed on the individual ΔFC matrices, thereby formally testing whether the T0-to-T1 change differed between stimulation conditions. Both directional contrasts were evaluated when appropriate. The same cluster-forming threshold and permutation procedure described above were applied, with condition labels randomly exchanged within subjects to preserve the repeated-measures structure. Significant components, therefore, represented connected subnetworks in which stimulation-induced FC changes differed between conditions.

The statistical analyses were performed using MATLAB 2025b and NBS toolbox. The level of significance was set at $p<0.05$. Graphs were done using custom-written scripts on MATLAB 2023b and R.

**References**

Conti M, D'Onofrio V, Bovenzi R, Ferrari V, Di Giuliano F, Cerroni R, et al. Cortical Functional Connectivity Changes in the Body-First and Brain-First Subtypes of Parkinson's Disease. Movement Disorders 2024. https://doi.org/10.1002/mds.30071.

Conti M, Guerra A, Pierantozzi M, Bovenzi R, D'Onofrio V, Simonetta C, et al. Band-Specific Altered Cortical Connectivity in Early Parkinson's Disease and its Clinical Correlates. Movement Disorders 2023:1–13. https://doi.org/10.1002/mds.29615.

De Micco R, Tessitore PA, Di Nardo F, De Mase A, Giordano A, Caiazzo G, et al. Sex-specific pattern of sensori-motor network connectivity in de novo Parkinson's disease patients. Eur J Neurol 2018;25.

Rossini PM, Burke D, Chen R, Cohen LG, Daskalakis Z, Di Iorio R, et al. Non-invasive electrical and magnetic stimulation of the brain, spinal cord, roots and peripheral nerves: Basic principles and procedures for routine clinical and research application: An updated report from an I.F.C.N. Committee. Clinical Neurophysiology 2015;126. https://doi.org/10.1016/j.clinph.2015.02.001.

Yassine S, Gschwandtner U, Auffret M, Achard S, Verin M, Fuhr P, et al. Functional Brain Dysconnectivity in Parkinson's Disease: A 5-Year Longitudinal Study. Movement Disorders 2022;37:1444–53. https://doi.org/10.1002/mds.29026.

Zalesky A, Fornito A, Bullmore ET. Network-based statistic: Identifying differences in brain networks. Neuroimage 2010;53. https://doi.org/10.1016/j.neuroimage.2010.06.041.
